# Ovarian carcinoma patients with a tumor *BRCA*-like genomic copy number aberration profile benefit from maintenance olaparib/bevacizumab therapy in the PAOLA-1 randomized controlled trial

**DOI:** 10.1101/2023.05.27.23290631

**Authors:** Philip C. Schouten, Sandra Schmidt, Kerstin Becker, Holger Thiele, Peter Nürnberg, Lisa Richters, Corinna Ernst, Isabelle Treilleux, Jacques Medioni, Florian Heitz, Carmela Pisano, Yolanda Garcia, Edgar Petru, Sakari Hietanen, Nicoletta Colombo, Ignace Vergote, Shoji Nagao, Sabine C. Linn, Eric Pujade Lauraine, Isabelle Ray-Coquard, Philip Harter, Eric Hahnen, Rita K. Schmutzler

## Abstract

**Background:** We previously established an ovarian carcinoma (OC) *BRCA*-like genomic copy number aberration profile classifier (“*BRCA*-like classifier”), which identifies tumors with deleterious mutations and epigenetic alterations in the homologous recombination pathway (1). We explored whether the classifier may also be predictive for therapies targeting tumors with homologous recombination deficiency (HRD) such as olaparib, a PARP inhibitor with synthetic lethal interaction with HRD, in combination with bevacizumab.

**Methods:** As part of the ENGOT (European Network of Gynaecological Oncological Trial groups) HRD initiative, the OC *BRCA*-like classifier was evaluated using tumor-derived DNA samples from 469 out of 806 patients enrolled in the PAOLA-1/ENGOT-ov25 trial. PAOLA-1 is a randomized, double-blind, international phase 3 trial (NCT02477644) including advanced high grade OC patients. Prolonged progression-free survival (PFS) and overall survival (OS) was observed for patients treated with maintenance olaparib and bevacizumab versus placebo and bevacizumab, and particularly for those patients tested HRD positive according to Myriad MyChoice® CDx HRD test.

**Results:** Results were obtained for 442 patients (failure rate of 6%, 27 of 469 samples). A survival benefit from adding maintenance olaparib was observed in the 298 (67%) patients with a *BRCA*-like tumor (hazard ratio (HR) for PFS: 0.49, 95% confidence interval (CI) 0.37-0.65, p = 0.01; OS: 0.64, 95% CI 0.45-0.90, p < 0.01). No benefit was detected in patients with a non-*BRCA*-like tumor when treated with olaparib (HR for PFS: 1.02, 95% CI 0.68-1.51, p = 0.93; OS 1.48, 95% CI 0.94-2.33, p = 0.09). P values for interaction between *BRCA*-like status and olaparib for PFS and OS were both 0.004. Multivariate analysis revealed comparable results. The concordance rate with the Myriad test was 77% in samples that were successfully analysed with both assays. In the survival analyses, the CIs of the *BRCA*-like classifier and the Myriad test overlap.

**Conclusion:** The *BRCA*-like classifier is a high-sensitive predictive biomarker for survival benefit of olaparib/bevacizumab as maintenance therapy in advanced ovarian carcinoma with a low drop-out rate.

## Introduction

First-line maintenance therapy with PARP inhibitor with bevacizumab was approved for advanced high grade ovarian carcinoma (OC) by the United States Food and Drug Association (FDA) and the European Medicines Agency (EMA) following the results of the PAOLA-1 trial (2). Mechanistically, PARP inhibitors induce synthetic lethality in patients with tumors harboring defects in homologous recombination, which are present in approximately 50% of high grade serous OC (3,4). In the PAOLA-1 trial improved survival was observed when olaparib was added to maintenance bevacizumab in patients with homologous recombination deficient (HRD) tumors, with or without tumor *BRCA1/2* mutations based on Myriad MyChoice® CDx, but not in patients with homologous recombination proficient tumors (5). A wide range of potential tests to identify HRD tumors are available, ranging from mutational testing, mutational and copy number signatures, to functional assays (6). However, there is a need for improvement of defining the patient subgroups, as globally there is a lack of consistently defined subgroups, a lack of negative predictive value in most of these tests and an inadequate assessment of the complex and dynamic nature of HRD phenotype (6). Despite these shortcomings, the EMA stated that HRD should be determined with a validated test by an experienced laboratory, without specifying the test (7). For research groups and molecular pathologists decentralized testing, in which the assay can be controlled within their own laboratory environment is an important benefit. Furthermore, health care systems do not (always) allow reimbursement for tests conducted in central laboratories abroad such as the Myriad MyChoice® CDx test, which is performed in the USA exclusively (8). Therefore, the ENGOT HRD initiative aimed to identify new reliable and feasible HRD tests, developed by European academic laboratories associated with the ENGOT trial groups (9).

We developed a *BRCA*-like genomic copy number aberration profile classifier (“*BRCA*-like classifier”) that predicts whether a tumor has similar aberrations as those observed in tumors from *BRCA*1 and *BRCA*2 germline mutant patients, so-called *BRCA*-like status (1). The signature is assumed to identify similar abnormalities as found in *BRCA*1/2-mutated tumors and may identify tumors that are HRD. When applied to the observational AGO-TR1 study, the classifier assigns *BRCA*-like status to tumors of patients with germline and somatic *BRCA*1 and *BRCA*2 mutations, along with *BRCA*1 promoter hypermethylation and pathogenic germline variants in *RAD51C*. Other HR pathway genes were not statistically assessable, possibly due to their low mutation prevalence in the cohort (1). When we applied our test to patients in the phase 3 OVHIPEC trial (NCT00426257), which randomized between addition of heated intraperitoneal chemotherapy (HIPEC) to standard neo-adjuvant carboplatin-paclitaxel or standard neoadjuvant carboplatin-paclitaxel treatment, we observed that patients with a non-*BRCA*-mutant HRD tumor benefited from the addition of HIPEC treatment, which served as preliminary evidence to predictive value (10). However, the use of HIPEC therapy in general and this trial have been debated extensively (11) and unexpectedly *BRCA* mutant patients did not benefit, likely due to a prognostic benefit (as of yet, of uncertain origin) (10).

In this investigation, we applied our test to a subgroup of 469 patients enrolled in the PAOLA-1 trial who had tumor DNA available for the present study, aiming to investigate whether it could predict survival benefit to olaparib maintenance therapy in 1^st^ line advanced OC.

## Material and Methods

### Setup of the ENGOT European HRD Initiative

The ENGOT European HRD Initiative consisted of three subsequent phases. In the first phase, putative tests beyond HR gene panel sequencing were identified and selected to proceed to phase 2. In phase 2, DNA samples of 85 tumors were available to assess predictive value of the test and to train/improve, if necessary. If the test appeared to be predictive, an additional 384 tumor DNA samples were analyzed in phase 3 of the program. For statistical analyses we combined the two cohorts because we did not need to perform any further training using the phase 2 cohort data.

### Samples and clinical data

The PAOLA-1 trial randomized 806 patients with newly diagnosed advanced (FIGO stage 3 or 4) high grade serous or endometrioid OC, primary peritoneal carcinoma, or fallopian tube carcinoma as previously described (12). Of the total trial population, 469 patients had sufficient tumor DNA sample available for the current initiative. We received 100 ng of DNA and the clinical data comprising original stratification factors, namely mutation status and response to first line treatment, age, performance status, timing of surgery, result of surgery, FIGO stage, histological subtype, Myriad MyChoice®CDx HRD test result, time to randomization, pre-treatment CA-125 measurement, progression free survival (PFS), PFS after second line treatment, and overall survival (OS). The trial was performed in accordance with the provisions of the Declaration of Helsinki and Good Clinical Practice guidelines under the auspices of an independent data monitoring committee and the patients provided written informed consent.

### Low coverage next-generation sequencing

The 85 phase 2 samples were sequenced at the Netherlands Cancer Institute (NKI) as described before (1). After validation of concordance between the NKI and Cologne Center for Genomics (CCG) sequencing facilities (Supplemental Materials), the phase 3 samples were sequenced at CCG as follows. A total of 384 DNA samples (100 ng each) isolated from formalin fixed paraffin embedded (FFPE) tumors were provided by ARCAGY Research (8 Rue Lamennais, 75008 Paris, France). All DNA samples were treated with NEBNext FFPE DNA Repair Mix (M6630L, New England Biolabs GmbH, Frankfurt am Main, Germany) according to the manufacturer’s protocol. Mechanical shearing of the DNA was performed using a Bioruptor NGS sonication system (UCD-600, Diagenode, 4102 Seraing (Ougrée), Belgium). DNA fragmentation was evaluated using the Tape Station 4200 (G2991BA, Agilent Technologies, Santa Clara, USA) and (high sensitivity) D1000 ScreenTapes (5067-5582, 5067-5584, Agilent). For library preparation, the TruSeq Nano DNA High Throughput Library Prep Kit (96 samples) was used (20015965, Illumina, San Diego, USA). The library pool was quantified by using the Peqlab KAPA Library Quantification Kit and the Applied Biosystems 7900HT Sequence Detection System and then sequenced on an Illumina NovaSeq 6000 sequencing instrument with a PE100 sequencing protocol (one SP flowcell).

### Quality control and BRCA-like classification

Copy number aberration profiles were generated by aligning the low coverage whole genome sequencing (WGS) reads to the hg38 reference genome, read counting in bins, and normalization for mappability and GC content within the dataset, and subsequently normalization to the reference training dataset. This input was used for the shrunken centroid classifier that classified the profile as *BRCA*-like (posterior probability > 0.5) or non-*BRCA*-like (posterior probability ≤ 0.5) as described before (1). Quality control was performed using noise variance and signal to noise ratio of the obtained copy number profiles in conjunction with visual assessment. The classifier will be made available on https://classifide.nki.nl.

### Statistics

Baseline characteristics were tabulated and assessed with respectively Fisher’s exact and chi-square tests. The Kaplan–Meier method was used to assess PFS (time from randomization until investigator-assessed disease progression or death), and OS (time from randomization until death). The log-rank test was used to assess the statistical significance of the differences in survival between the olaparib and the placebo group. We used Cox regression analysis to estimate the hazard ratio (HR), 95% confidence interval (CI), p value and p value for interaction comparing the olaparib and placebo group. For multivariate analyses we used stratification factors, as they appeared to capture best confounding effects from the available data. The analyses were performed in the *BRCA*-like and non-*BRCA*-like group. All tests were two-sided with p values < 0.05 being considered significant.

## Results

We pooled the test results from phases 2 and 3 of the ENGOT HRD project (n=85 and n=384, total n=469), as we did not perform any training or optimization of the model during phase 2 of this study. Eight samples failed sequencing. An additional 19 samples were excluded by a combined statistical and visual quality control of the copy number aberration profile. For 3 of these, noisy profiles were obtained, and for the further 16, resulting profiles showed few or low-level aberrations, indicating a low(er than estimated) tumor cell content.

Table 1 describes the cohort characteristics, showing balanced subgroups for histology, FIGO stage and first line therapy response. Pathogenic variants in *BRCA*1 and *BRCA*2 were enriched in patients with *BRCA*-like tumors. All other clinicopathological characteristics appeared evenly distributed across patients with *BRCA*-like and non-*BRCA*-like tumors.

**Table 1.**
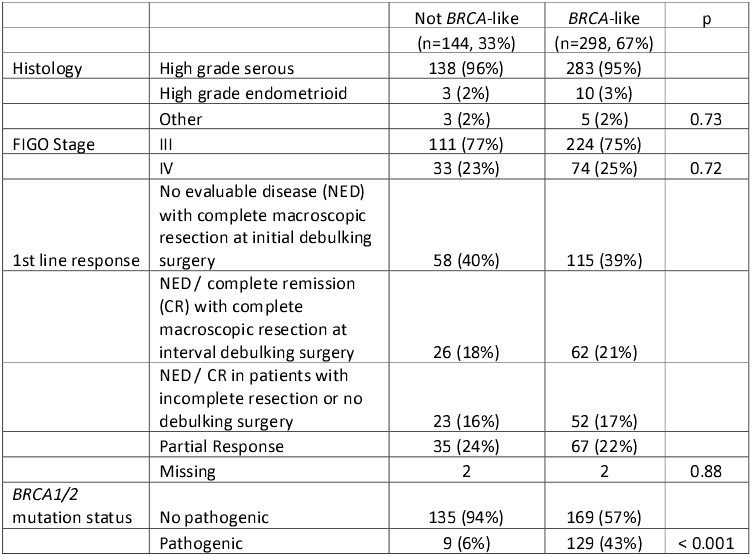
Patient characteristics Patient characteristics stratified by *BRCA*-like tumor status.

Figure 1, A-D shows the Kaplan-Meier survival analysis of patients with *BRCA*-like tumors and non-*BRCA*-like tumors, comparing olaparib plus bevacizumab vs placebo plus bevacizumab treatment for PFS and OS, and Table 2 shows the associated HRs. Considering PFS, for patients with non-*BRCA*-like tumors similar survival was observed for experimental arm and control arm (median PFS 17.6 vs 16.6 months, HR: 1.02, 95% CI 0.68-1.51), whereas patients with a *BRCA*-like tumor survived longer on average on treatment with olaparib and bevacizumab (PFS 36.4 vs 18.6 months HR 0.49 (95% CI: 0.37-0.65, p < 0.001). For patients with non-*BRCA*-like tumors, OS did not significantly differ between both treatment arms, however there may be a trend for poorer survival on olaparib plus bevacizumab (38.8 vs 52.3 months, HR: 1.48, 95% CI: 0.94-2.33, p = 0.09). Patients with *BRCA*-like tumors showed longer survival after olaparib plus bevacizumab (75.2 vs 54.9 months, HR 0.64, 95%CI 0.45-0.90, p:0.01). P values retrieved for interaction between *BRCA*-like status and treatment were both 0.004. *BRCA*-like status did not confer a prognostic survival benefit. Supplemental Figure 6 shows Kaplan-Meier curves split out between patients with *BRCA*-like and mutant, *BRCA*-like non-mutant, non-*BRCA*-like mutant and non-*BRCA*-like non-mutant tumors, the curves and interpretations resemble those presented in Figure 1.

**Table 2.**
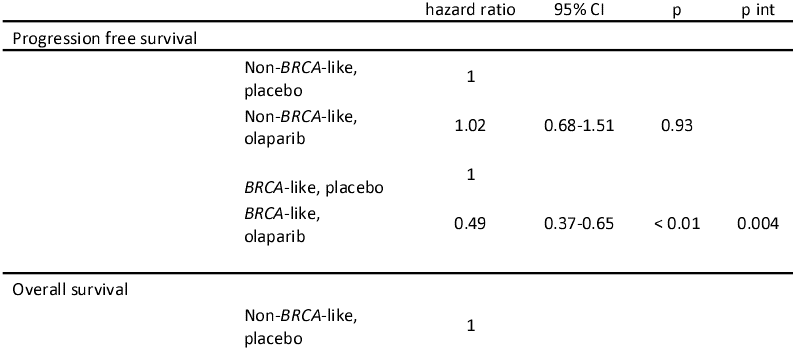
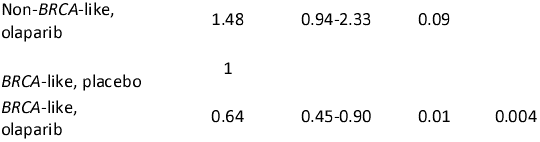
Cox regression analysis results considering progression free survival (PFS) and overall survival (OS), olaparib vs placebo, in patients with non-*BRCA*-like and *BRCA*-like tumors. CI = confidence interval, p int = p value for interaction between *BRCA*-like and treatment arm. The placebo arm is the reference (HR = 1).

|  |  | hazard ratio | 95% CI | p | p int |
| --- | --- | --- | --- | --- | --- |
| Progression free survival |  |  |  |  |  |
|  | Non-*BRCA*-like, placebo | 1 |  |  |  |
|  | Non-*BRCA*-like, olaparib | 1.02 | 0.68-1.51 | 0.93 |  |
|  | *BRCA*-like, placebo | 1 |  |  |  |
|  | *BRCA*-like, olaparib | 0.49 | 0.37-0.65 | < 0.01 | 0.004 |
| Overall survival |  |  |  |  |  |
|  | Non-*BRCA*-like, placebo | 1 |  |  |  |

Table 2. Cox regression analysis results considering progression free survival (PFS) and overall survival (OS), olaparib vs placebo, in patients with non-*BRCA*-like and *BRCA*-like tumors.
|  |  |  |  |  |
| --- | --- | --- | --- | --- |
| Non-*BRCA*-like,<br>olaparib | 1.48 | 0.94-2.33 | 0.09 |  |
| *BRCA*-like, placebo | 1 |  |  |  |
| *BRCA*-like,<br>olaparib | 0.64 | 0.45-0.90 | 0.01 | 0.004 |

**Figure 1.**
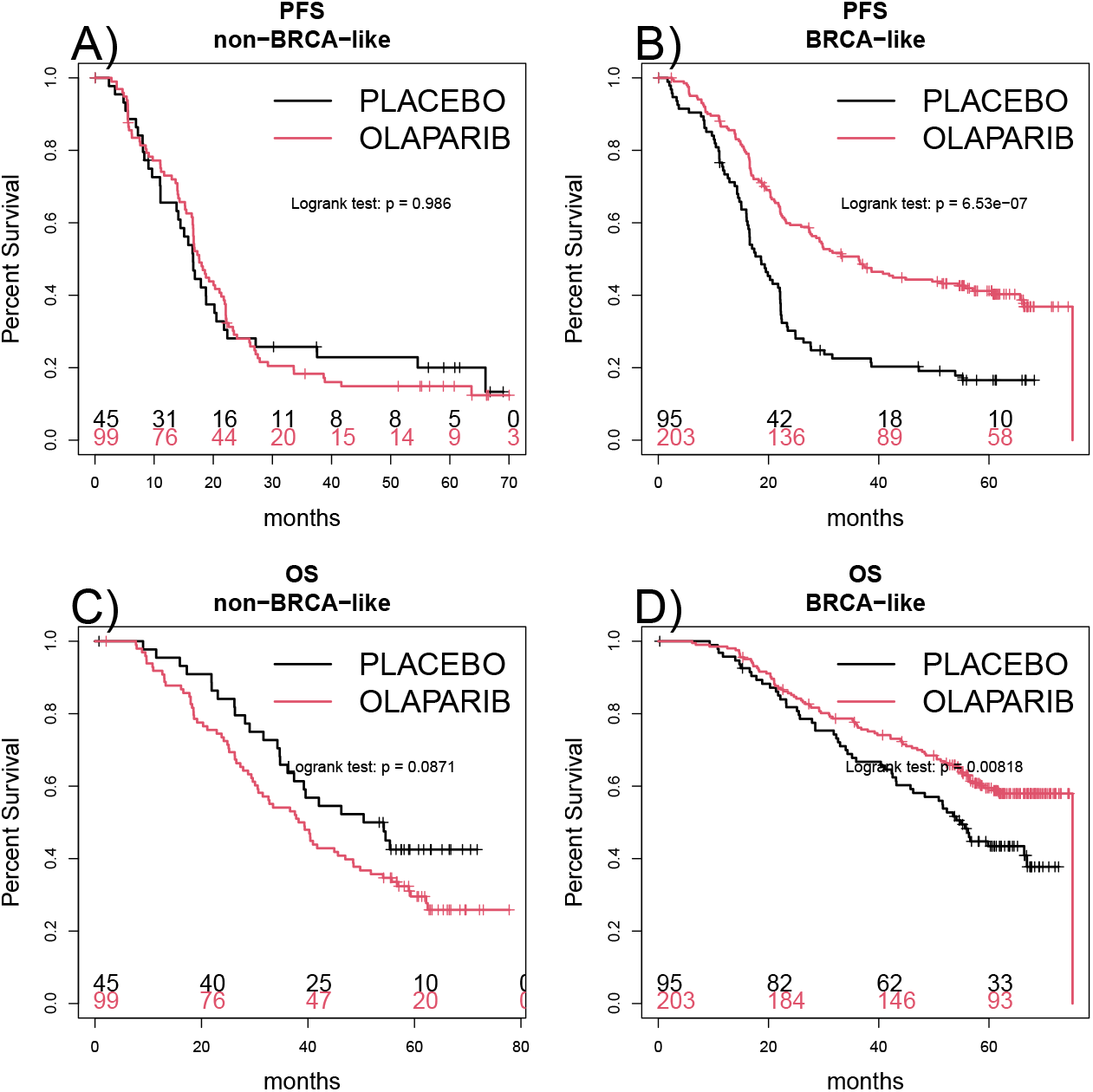
Kaplan-Meier curves. Progression free survival (PFS) of patients with non-*BRCA*-like tumors (A) and patients with *BRCA*-like (B) tumors treated with olaparib plus bevacizumab or placebo plus bevacizumab, respectively. Overall survival (OS) of patients with non-*BRCA*-like tumors (C) and patients with *BRCA*-like tumors (D) treated with olaparib plus bevacizumab or placebo plus bevacizumab.

These observations remained in multivariate analyses (Table 3). Response on first line therapy was an independent prognostic variable. For PFS, patients with *BRCA*-like tumors benefitted from olaparib (HR 0.46, 95% CI 0.34-0.61, p < 0.001). Patients with non-*BRCA*-like tumors did not benefit from olaparib, (HR 1.005, 95%CI 0.67-1.50, p = 0.98; p value for interaction = 0.002). For OS, patients with *BRCA*-like tumors benefitted, (HR 0.62, 95%CI 0.44-0.87, p=0.006, whereas for patients with non-*BRCA*-like tumors an increased HR was observed (1.51, 95%CI 0.95-2.40, p=0.08, p value for interaction = 0.002).

**Table 3.** Multivariate Cox proportional hazards models for progression free survival and overall survival to estimate the hazard ratio (HR) of olaparib plus bevacizumab vs placebo plus bevacizumab treatment in patients with non-*BRCA*-like tumors and *BRCA*-like tumors. NED = no evaluable disease, CR = complete remission. CI = confidence interval, p int = p value for interaction between *BRCA*-like and treatment arm. The placebo arm is the reference (HR = 1).

|  |  |  | HR | 95% CI | p | p int |
| --- | --- | --- | --- | --- | --- | --- |
| Progression free survival |  |  |  |  |  |  |
|  | Response 1st<br>line treatment |  |  |  |  |  |
|  |  | NED with complete<br>macroscopic resection<br>at initial debulking<br>surgery | 1.00 |  |  |  |
|  |  | NED / CR with<br>complete macroscopic<br>resection at interval<br>debulking surgery | 1.85 | 1.34-2.54 | < 0.01 |  |
|  |  | NED / CR in patients<br>with incomplete<br>resection or no<br>debulking surgery | 2.18 | 1.56-3.05 | < 0.01 |  |
|  |  | Partial Response | 2.52 | 1.87-3.40 | < 0.01 |  |
|  | FIGO stage | III | 1.00 |  |  |  |
|  |  | IV | 1.26 | 0.97-1.63 | 0.09 |  |
|  | *BRCA*-like |  | 0.93 | 0.62-1.40 | 0.74 |  |
|  | Non-*BRCA*-like,<br>olaparib |  | 1.00 | 0.67-1.50 | 0.98 |  |
|  | *BRCA*-like,<br>olaparib |  | 0.46 | 0.34-0.61 | < 0.01 | 0.002 |
| Overall survival |  |  |  |  |  |  |
|  | Response 1st<br>line treatment |  |  |  |  |  |
|  |  | NED with complete<br>macroscopic resection<br>at initial debulking<br>surgery | 1.00 |  |  |  |
|  |  | NED / CR with<br>complete macroscopic<br>resection at interval<br>debulking surgery | 2.28 | 1.53-3.39 | < 0.01 |  |
|  |  | NED / CR in patients<br>with incomplete<br>resection or no<br>debulking surgery | 3.22 | 2.17-4.78 | < 0.01 |  |
|  |  | Partial Response | 3.70 | 2.58-5.32 | < 0.01 |  |
|  | FIGO stage | III | 1.00 |  |  |  |
|  |  | IV | 1.04 | 0.78-1.40 | 0.78 |  |
|  | *BRCA*-like |  | 0.95 | 0.59-1.54 | 0.84 |  |
|  | Non-*BRCA*-like,<br>olaparib |  | 1.51 | 0.95-2.40 | 0.08 |  |
|  | *BRCA*-like,<br>olaparib |  | 0.62 | 0.44-0.87 | 0.01 | 0.002 |

### Comparison with Myriad MyChoice®CDx

The *BRCA*-like classifier analysis failed in 27/469 cases (5.7%), and the MyChoice test failed in 44/469 cases (9.4 %). Only 7 cases failed in both assays.

For a direct comparison, 405 samples were successfully analysed using both assays. A flow diagram of classification by both tests is shown in Supplemental Figure 7. Concordant results were obtained for 107 HRD/*BRCA*-like negative and 207 HRD/*BRCA*-like positive (n=314/405, 77.5%) tumors. Of the remaining tumors, 22 were Myriad MyChoice positive and non-*BRCA*-like, and 69 tumors were classified as *BRCA*-like Myriad MyChoice negative. Figure 2 shows the Kaplan-Meier curves of the subset of experiments that were successful for both tests, and corresponding HRs are listed in Table 4. The Kaplan-Meier curves and HRs were comparable, and the confidence intervals overlap. The overall effects reflected the analyses in the full cohort. Despite more patients having a *BRCA*-like tumor, no deleterious effect was observed with respect to the interaction between the *BRCA*-like classifier and olaparib treatment, which remained statistically significant.

**Table 4.** Hazard ratios, 95% Confidence intervals and p-values comparing progression free survival (PFS) and overall survival (OS) for olaparib plus bevacizumab vs. placebo plus bevacizumab treatment of patients with biomarker positive and negative tumors.

| PFS |  | HR | 95%CI | p |
| --- | --- | --- | --- | --- |
|  | Myriad negative placebo | 1.00 |  |  |
|  | Myriad negative olaparib | 1.04 | 0.72-1.49 | 0.84 |
|  | non-*BRCA*-like placebo | 1.00 |  |  |
|  | non-*BRCA*-like olaparib | 1.00 | 0.65-1.55 | 0.99 |
|  | Myriad positive placebo | 1.00 |  |  |
|  | Myriad positive olaparib | 0.38 | 0.26-0.54 | < 0.01 |
|  | *BRCA*-like placebo | 1.00 |  |  |
|  | *BRCA*-like olaparib | 0.48 | 0.35-0.65 | <0.01 |
| OS |  |  |  |  |
|  | Myriad negative placebo | 1.00 |  |  |
|  | Myriad negative olaparib | 1.27 | 0.85-1.9 | 0.24 |
|  | non-*BRCA*-like placebo | 1.00 |  |  |
|  | non-*BRCA*-like olaparib | 1.54 | 0.93-2.57 | 0.10 |
|  | Myriad positive placebo | 1.00 |  |  |
|  | Myriad positive olaparib | 0.55 | 0.35-0.85 | 0.01 |
|  | *BRCA*-like placebo | 1.00 |  |  |
|  | *BRCA*-like olaparib | 0.63 | 0.44-0.91 | 0.01 |

**Figure 2.**
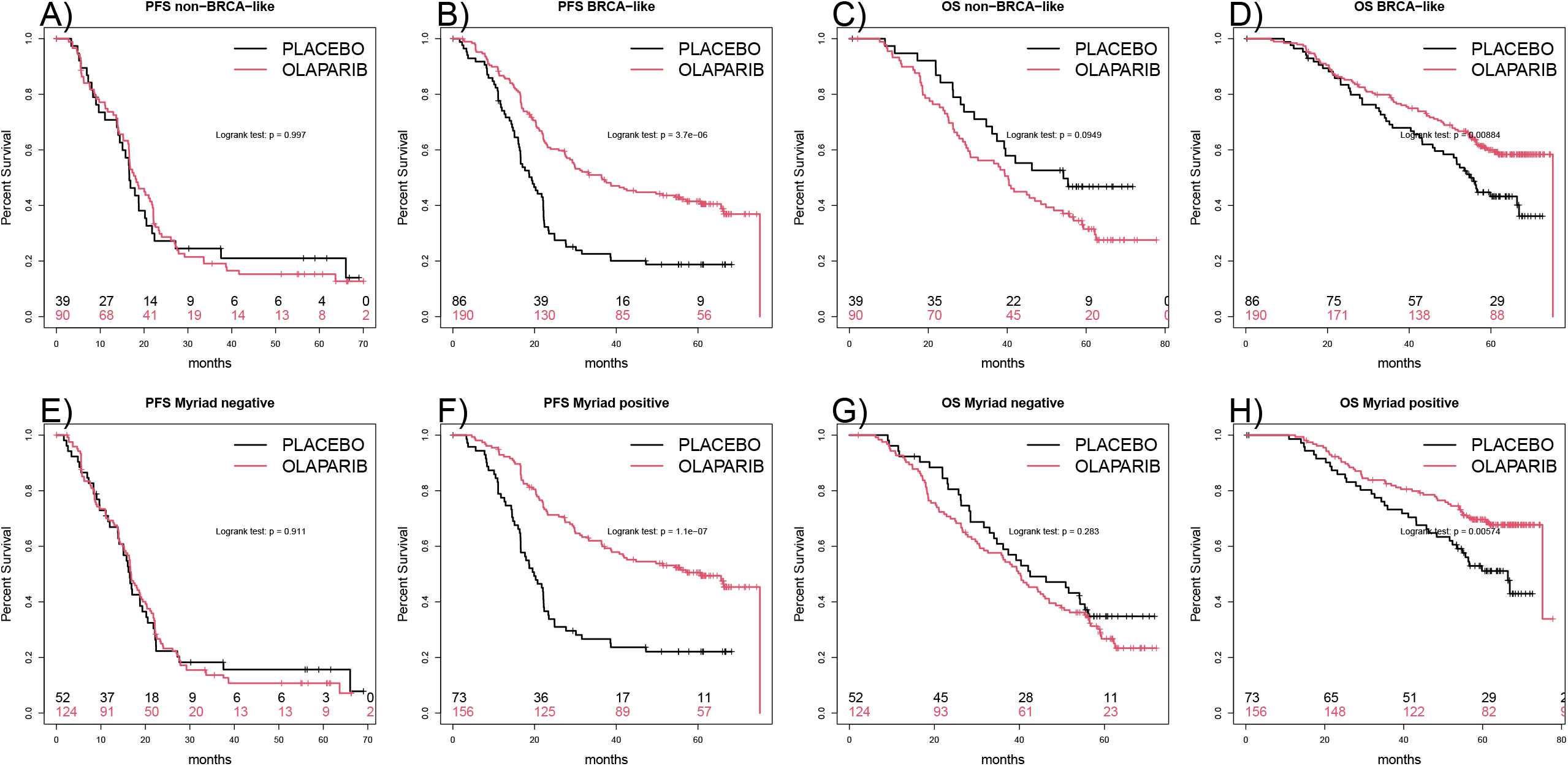
Kaplan-Meier curves for the subset of experiments that were successful for both tests. Progression free survival (PFS) of patients with non-*BRCA*-like tumors (A) and patients with *BRCA*-like tumors (B) treated with olaparib plus bevacizumab or placebo plus bevacizumab, respectively. Overall survival (OS) of patients with non-*BRCA*-like tumors (C) and patients with *BRCA*-like tumors (D) treated with olaparib plus bevacizumab or placebo plus bevacizumab. PFS of patients with Myriad myChoice negative tumors (E) and patients with Myriad myChoice positive tumors (F) treated with olaparib plus bevacizumab or placebo plus bevacizumab, respectively. OS of patients with Myriad myChoice negative tumors (G) and patients Myriad myChoice positive tumors (H) treated with olaparib plus bevacizumab or placebo plus bevacizumab.

Analysis of group of 91 samples with discordant test outcome, resulted inKaplan-Meier curves shown in Figure 3 and Cox proportional hazards analysis results shown in Table 5. Considering PFS, patients with Myriad MyChoice positive tumors showed statistically significant longer survival with olaparib than with placebo treatment (p < 0.01). For patients with a *BRCA*-like tumor, the difference was not statistically significant (p = 0.08). Considering OS, there was no survival difference between olaparib and placebo treatment for both tests. With respect to interpretation, it must be stressed that the subsets are small, that in these specific subsets there is a trend for longer survival on olaparib across all subgroups, the OS results do not follow the PFS results (contrary to the full cohort analyses).

**Table 5.** Results of multivariate Cox regression analyses with progression free survival (PFS), respectively overall survival (OS) as the outcome, for the subgroup of 91 samples with discordant test results. HR = hazard ratio, CI = confidence interval.

| Table 5.<br>PFS |  | HR | 95%CI | p |
| --- | --- | --- | --- | --- |
|  | Myriad negative/*BRCA*-like placebo | 1.00 |  |  |
|  | Myriad negative/*BRCA*-like olaparib | 0.59 | 0.59-1.06 | 0.08 |
|  | Myriad positive/non-*BRCA*-like placebo | 1.00 |  |  |
|  | Myriad positive/non-*BRCA*-like | 0.15 | 0.15- | < 0.01 |
|  | olaparib |  | 0.44 |  |
| OS |  |  |  |  |
|  | Myriad negative/*BRCA*-like placebo | 1.00 |  |  |
|  | Myriad negative/*BRCA*-like olaparib | 0.86 | 0.86-1.56 | 0.62 |
|  | Myriad positive/non-*BRCA*-like placebo | 1.00 |  |  |
|  | Myriad positive/non-*BRCA*-like olaparib | 0.67 | 0.67-2.41 | 0.54 |

**Figure 3.**
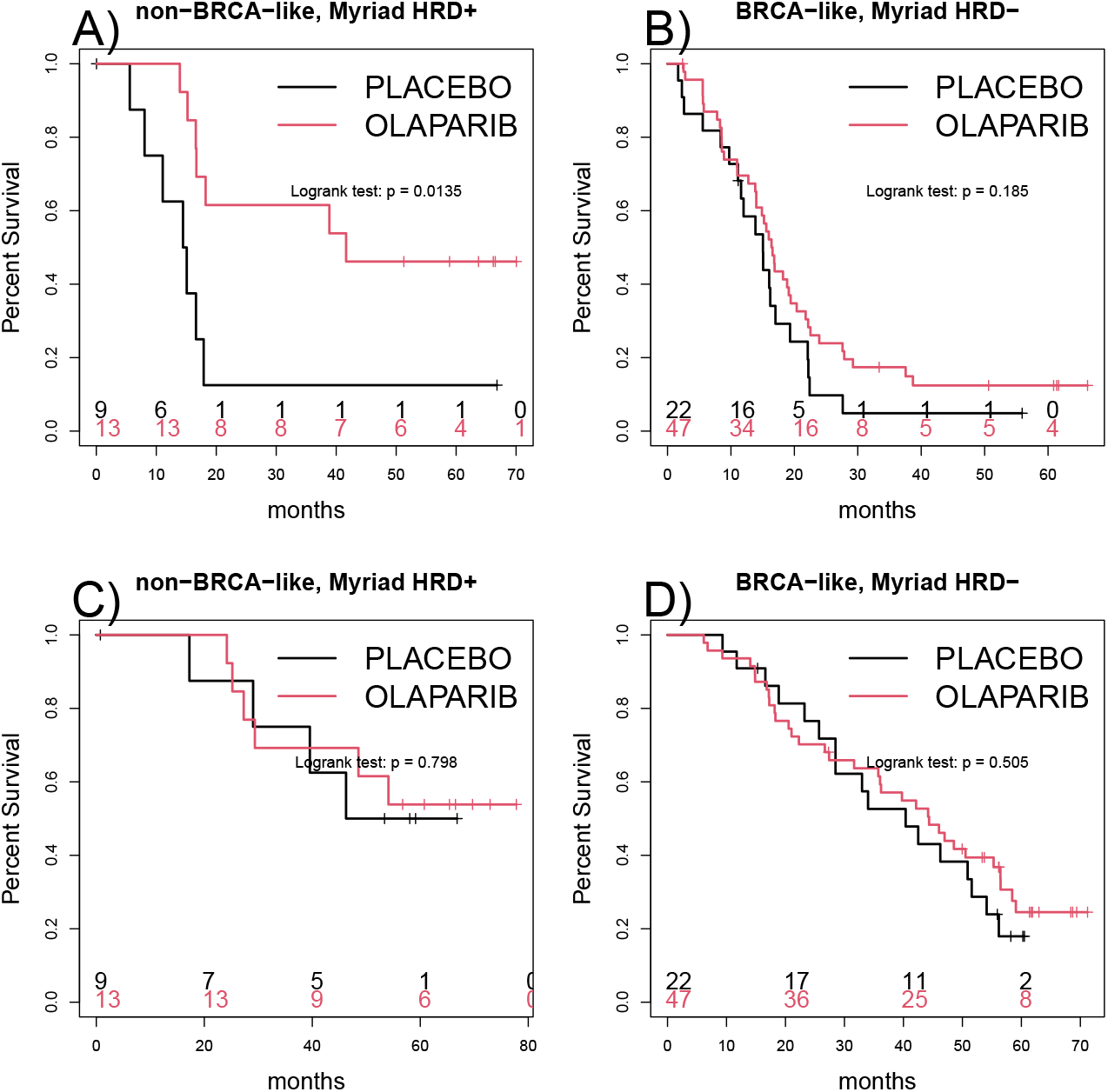
Kaplan-Meier curves of progression free survival (PFS, A and B), overall survival (OS, C and D) of patients that have a discordant conclusion between the two tests. Corresponding hazard ratios are shown in Table 5.

## Discussion

We investigated in this exploratory study whether the previously published *BRCA*-like tumor copy number aberration profile classifier (1) could be a predictor of efficacy for olaparib as maintenance therapy in advanced OC. We showed that patients with a tumor with *BRCA*-like signature had a survival benefit from the addition of olaparib to bevacizumab maintenance, whereas those with a tumor with a non-*BRCA*-like signature did not.

The PAOLA-1 phase 3 trial offered a unique opportunity to further investigate and validate academically new biomarkers for HRD at the European level (9,13–15). The two-step study design of the HRD initiative and the randomized nature of the trial generates high level of evidence for those biomarkers that showed predictive value (16,17), while using preciously rare DNA samples in a responsible manner. Therefore, it can serve as an example for future efforts aiming to identify new biomarkers.

Our *BRCA*-like signature as predictor of olaparib efficacy when added to bevacizumab maintenance in first-line therapy, was thus validated within a large group of patients with advanced high grade OC. In the current study, non-*BRCA*-like tumor status showed a non-significant trend for worse OS outcome after olaparib treatment of affected patients. Several hypotheses can be proposed to explain this observation. In the absence of HRD, continued treatment with an underperforming cytotoxic agent such as olaparib of patients with a non-*BRCA*-like tumor may induce further changes that promote tumor progression or resistance, which ultimately lead to decreased OS. This hypothesis has already been generated when a lower efficacy of subsequent treatments was observed after maintenance olaparib therapy compared to placebo even in patients with *BRCA* mutated tumor and thus with HRD (18). Another hypothesis is that the patients with non-*BRCA*-like tumors have had unspecific responses to the first-line treatment, and that the continued treatment with a non-targeted but cytotoxic agent (olaparib) delays more effective treatment. Knowing the HRD tumor status of the patients that did not respond to the first-line therapy would give additional insight but was not assessed. Arguably, response to platinum-based chemotherapy used as an inclusion criterion in PAOLA-1 trial may be considered as a surrogate marker for HRD. Since platinum is also targeting HRD, one can raise the question whether knowing the HRD status as early as possible would be decisive to choose not only the best candidates for PARP inhibitor maintenance therapy, but also the right chemotherapy treatment and to delineate a clear group for which current regimen might be suboptimal. We conclude that the *BRCA*-like signature may be effective in detecting those patients without benefit from the addition of olaparib, or even possibly may be even harmed.

The *BRCA*-like classifier can select a larger group of patients eligible for olaparib than the HRD group according to the Myriad myChoice assay. An additional advantage of our assay is the low drop-out rate, which could even be decreased further in a controlled laboratory setting and provision of percentage of viable tumor cells in the selected area, the ability to check the actual dissected region and/or the availability of variant allele fractions to derive tumor cell content.

Within our study we demonstrated that the assay can be performed in a decentralized way, which facilitates easy implementation in current workflows. Previously, copy number aberration data was demonstrated to be highly robust, and hence, it is expected that the assay can reliably be read out with a multitude of techniques although for clinical use specific validation studies are required (20). In addition, it should be noted that the *BRCA*-like classifier was developed independently of the current study. Thus, we did not perform any training on the PAOLA-1 cohort that might have biased the outcomes of the test towards the specific characteristics of the patients included in this trial. The ability of the *BRCA*-like classifier to predict the patients who benefit from heated intra-peritoneal chemotherapy (10) suggests that this test can be used outside of the specific setting of the PAOLA-1 trial.

We took the Myriad MyChoice CDx HRD test as a gold standard and comparing to this standard may lead to difficult to interpret situations, specifically when the concordance is moderate, as it was the case in our study. However, the predictive value of both tests was similar, with largely overlapping confidence intervals in the head-to-head comparisons restricted to the analyses of samples that had both biomarkers successfully measured. The analysis of samples with discordant test results was difficult to interpret due to the small size of the corresponding subgroups. Moreover, both unexpected patterns between various survival readouts and a tendency for benefit of olaparib in all patients in this subgroup were observed. Nevertheless, the comparison of the Kaplan-Meier curves suggests that olaparib compared to placebo may benefit to some of the patients who had *BRCA*-like tumors and had Myriad MyChoice HRD CDx negative tumors, and conversely.

With the EMA guidance that HRD testing can be performed with a validated test in an experienced laboratory, it happened that many other potential HRD biomarkers have been devised. The European ENGOT HRD initiative has allowed to validate several different tests developed in the academic setting (9,13–15). The comparison between these DNA-based HRD tests is challenging in particular when additional factors are considered such as practical/logistical issues (e.g. required amount of DNA, turn-around-times, access to algorithms, required equipments). In this context, the *BRCA*-like classifier is one of the potential tests to identify patients with HRD tumors. Considered entities and methodologies of existing approaches for tumor HRD testing range from mutations, genomic scar signatures, mutational signatures to functional tests (6). All these tests have shown to correlate with mechanistic explanations of HRD, such as underlying *BRCA* mutations, and responses to HRD-targeted therapy. Despite this, some of the biomarkers may represent a history of HRD-driven tumorigenesis, rather than actual HRD status, as it may have changed during tumor evolution (21). Functional assays may assess the actual status of the HR pathway (6,21,22) but the need of fresh tumor tissue limits their routine use. Interestingly, it has been suggested that genomic scars and functional read-outs may identify small non-overlapping groups (23). These observations suggest that the optimal HRD test is not yet available and prompt further research to develop and validate tests to detect HRD with increasing accuracy.

In conclusion, the *BRCA*-like classifier that has been previously established as HRD test, was validated on the current PAOLA-1 cohort as a predictive biomarker to select the patients who will most benefit from olaparib and bevacizumab maintenance in first-line treatment of advanced high grade ovarian carcinoma. It had a lower drop-out rate and resulted in a larger group of patients with biomarker-positive tumors than the reference HRD test and showed a trend towards poorer OS of patients with non-*BRCA*-like tumors treated with olaparib. Thus, it is a useful biomarker which can be applied in clinical routine practice. We are currently using prospectively the *BRCA*-like classifier in the AGO-OVAR28 study [NCT05009082].

## Data Availability

All data produced in the present study are available upon reasonable request to the authors, contingent on signing data transfer agreements.

## Funding

Genetic analyses were supported by the Köln Fortune Program, Faculty of Medicine, University of Cologne, Germany (#5/2022). The funding source had no role in the design and conduct of the study; collection, management, analysis, and interpretation of the data; preparation, review, or approval of the manuscript; nor in the decision to submit the manuscript for publication.

## Conflict of interest

PCS, SCL, EH and RKS are named inventors on the patent application “Methods for assessing homologous recombination deficiency in ovarian cancer cells, WO2023031121, international publication date 09.03.2023)

## Notes

### Funding Statement

Genetic analyses were supported by the Koln Fortune Program, Faculty of Medicine, University of Cologne, Germany (#5/2022). The funding source had no role in the design and conduct of the study; collection, management, analysis, and interpretation of the data; preparation, review, or approval of the manuscript; nor in the decision to submit the manuscript for publication.

### Author Declarations

The Ethic Committee "Comite de Protection des Personnes SUD-EST IV" of Centre Leon Berard, 28 rue Laennec, 69373 Lyon gave ethical approval for this work.

## References

1. Schouten PC, Richters L, Vis DJ, Kommoss S, van Dijk E, Ernst C, et al. Ovarian Cancer-Specific BRCA-like Copy-Number Aberration Classifiers Detect Mutations Associated with Homologous Recombination Deficiency in the AGO-TR1 Trial. Clin Cancer Res. 2021 Dec 1;27(23):6559–69.

2. Le Saux O, Vanacker H, Guermazi F, Carbonnaux M, Roméo C, Larrouquère L, et al. Poly(ADPribose) polymerase inhibitors in combination with anti-angiogenic agents for the treatment of advanced ovarian cancer. Future Oncol. 2021 Jun;17(18):2291–304.

3. Lord CJ, Ashworth A. BRCAness revisited. Nat Rev Cancer. 2016 Feb;16(2):110–20.

4. Bell D, Berchuck A, Birrer M, Chien J, Cramer DW, Dao F, et al. Integrated genomic analyses of ovarian carcinoma. Nature. 2011;474(7353):609–15.

5. Harter P, Mouret-Reynier MA, Pignata S, Cropet C, González-Martín A, Bogner G, et al. Efficacy of maintenance olaparib plus bevacizumab according to clinical risk in patients with newly diagnosed, advanced ovarian cancer in the phase III PAOLA-1/ENGOT-ov25 trial. Gynecol Oncol. 2022 Feb;164(2):254–64.

6. Miller RE, Leary A, Scott CL, Serra V, Lord CJ, Bowtell D, et al. ESMO recommendations on predictive biomarker testing for homologous recombination deficiency and PARP inhibitor benefit in ovarian cancer. Annals of Oncology. 2020 Dec;31(12):1606–22.

7. EMA. Lynparza - EPAR - product information - EMEA/H/C/003726 - II/0053 [Internet]. [cited 2023 Mar 16]. Available from: https://www.ema.europa.eu/en/documents/product-information/lynparza-epar-product-information_en.pdf

8. Denkert C, Romey M, Swedlund B, Hattesohl A, Teply-Szymanski J, Kommoss S, et al. Homologous Recombination Deficiency as an Ovarian Cancer Biomarker in a Real-World Cohort: Validation of Decentralized Genomic Profiling. J Mol Diagn. 2022 Dec;24(12):1254–63.

9. Pujade-Lauraine E, Christinat Y, D’incalci M, Schouten P, Buisson A, Heukamp L, et al. 201 Homologous recombination deficiency testing in advanced ovarian cancer: description of the ENGOT HRD European initiative. International Journal of Gynecologic Cancer 2021;31:A208.

10. Koole SN, Schouten PC, Hauke J, Kluin RJC, Nederlof P, Richters LK, et al. Effect of HIPEC according to HRD/BRCAwt genomic profile in stage III ovarian cancer - results from the phase III OVHIPEC trial. Int J Cancer. 2022 May 18;

11. Harter P, Bogner G, Chiva L, Cibula D, Concin N, Fotopoulou C, et al. Statement of the AGO Kommission Ovar, AGO Study Group, NOGGO, AGO Austria, Swiss AGO, BGOG, CEEGOG, GEICO, and SFOG regarding the use of hyperthermic intraperitoneal chemotherapy (HIPEC) in epithelial ovarian cancer. Bull Cancer. 2023 Mar 24;S0007-4551(23)00098-X.

12. Ray-Coquard I, Pautier P, Pignata S, Pérol D, González-Martín A, Berger R, et al. Olaparib plus Bevacizumab as First-Line Maintenance in Ovarian Cancer. N Engl J Med. 2019 Dec 19;381(25):2416–28.

13. Willing EM, Vollbrecht C, Voessing C, Weist P, Schallenberg S, Jori B, et al. 2022-RA-873-ESGO Validation study of the ‘NOGGO-GIS ASSAY’ based on ovarian cancer samples from the first-line PAOLA-1/ENGOT-ov25 phase-III trial. In: Pathology [Internet]. BMJ Publishing Group Ltd; 2022 [cited 2023 Mar 16]. p. A370.1-A370. Available from: https://ijgc.bmj.com/lookup/doi/10.1136/ijgc-2022-ESGO.791

14. Loverix L, Vergote I, Busschaert P, Vanderstichele A, Boeckx B, Venken T, et al. Predictive value of the Leuven HRD test compared with Myriad myChoice PLUS on 468 ovarian cancer samples from the PAOLA-1/ENGOT-ov25 trial (LBA 6). Gynecologic Oncology. 2022 Aug;166:S51–2.

15. Christinat Y, Ho L, Clément S, Genestie C, Sehouli J, Martin AG, et al. 2022-RA-567-ESGO The Geneva HRD test: clinical validation on 469 samples from the PAOLA-1 trial International Journal of Gynecologic Cancer 2022;32:A238–A239.

16. Simon RM, Paik S, Hayes DF. Use of archived specimens in evaluation of prognostic and predictive biomarkers. J Natl Cancer Inst. 2009 Nov 4;101(21):1446–52.

17. Sargent DJ, Mandrekar SJ. Statistical issues in the validation of prognostic, predictive, and surrogate biomarkers. Clin Trials. 2013 Oct;10(5):647–52.

18. Frenel JS, Kim JW, Aryal N, Asher R, Berton D, Vidal L, et al. Efficacy of subsequent chemotherapy for patients with BRCA1/2-mutated recurrent epithelial ovarian cancer progressing on olaparib versus placebo maintenance: post-hoc analyses of the SOLO2/ENGOT Ov-21 trial. Ann Oncol. 2022 Oct;33(10):1021–8.

19. Muggia FM, Braly PS, Brady MF, Sutton G, Niemann TH, Lentz SL, et al. Phase III randomized study of cisplatin versus paclitaxel versus cisplatin and paclitaxel in patients with suboptimal stage III or IV ovarian cancer: a gynecologic oncology group study. J Clin Oncol. 2000 Jan;18(1):106–15.

20. Schouten PC, Grigoriadis A, Kuilman T, Mirza H, Watkins JA, Cooke SA, et al. Robust BRCA1-like classification of copy number profiles of samples repeated across different datasets and platforms. Mol Oncol. 2015 Aug;9(7):1274–86.

21. Doig KD, Fellowes AP, Fox SB. Homologous Recombination Repair Deficiency: An Overview for Pathologists. Mod Pathol. 2023 Jan 10;36(3):100049.

22. Stewart MD, Merino Vega D, Arend RC, Baden JF, Barbash O, Beaubier N, et al. Homologous Recombination Deficiency: Concepts, Definitions, and Assays. The Oncologist. 2022 Mar 11;27(3):167–74.

23. Meijer TG, Nguyen L, Van Hoeck A, Sieuwerts AM, Verkaik NS, Ladan MM, et al. Functional RECAP (REpair CAPacity) assay identifies homologous recombination deficiency undetected by DNA-based BRCAness tests. Oncogene. 2022 Jun;41(26):3498–506.

