## Supplemental Methods and Figures for "Ovarian carcinoma patients with a tumor *BRCA*-like genomic copy number aberration profile benefit from maintenance olaparib/bevacizumab therapy in the PAOLA-1 randomized controlled trial"

**Supplementary methods**

*Proof of concept: decentralized distribution*

Previously, we employed a HiSeq2500 sequencer with 65bp read length at the Netherlands Cancer Institute (NKI) (1). As proof of concept of decentralized distribution and to investigate robustness of the assay, we conducted sequencing at the Cologne Center for Genomics (CCG) of the Phase 3 samples. For higher throughput we employed Illumina NovaSeq6000 sequencer. We sequenced 47 paired samples from the AGOTR1 study with the NovaSeq machine (see Materials and Methods) and compared the results of classification with those previously obtained with the HiSeq machine(1). Read-outs for successful validation comprised similarity of the (average) profiles between both facilities/techniques and concordance of classification.

We plotted average log ratio and segmented log ratio copy number profiles of the samples sequenced at NKI and CCG. Visual inspection confirmed that the average profiles overlap, see Supplementary Figure 1.

Supplementary Figure 1. Copy number aberration profiles of 47 samples sequenced at NKI and CCG.

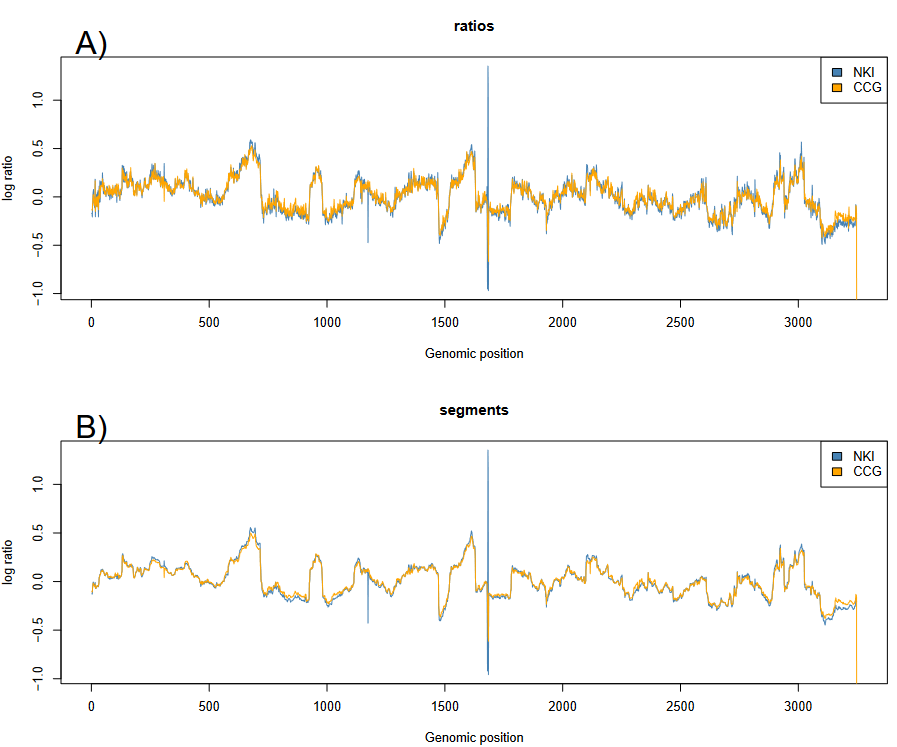

Supplementary Figure 1. Average unsegmented (A) and segmented (B) copy number profiles of 47 samples that were sequenced at NKI (blue) and CCG (orange).

When we plotted the sorted log ratios and sorted segmented log ratios, there was a strong correlation between the two platforms, with the (segmented) log ratios mostly on the identity line of the plot (Supplementary Figure 2). Note, this measure resembles quantile (normalized) plots of the distribution and not the actual distribution or correlation between the profiles, i.e. range and centering of (segmented) log ratios obtained by the two sequencing facilities.

Supplementary Figure 2. Correlation of the distribution between 47 samples sequenced at NKI and CCG.

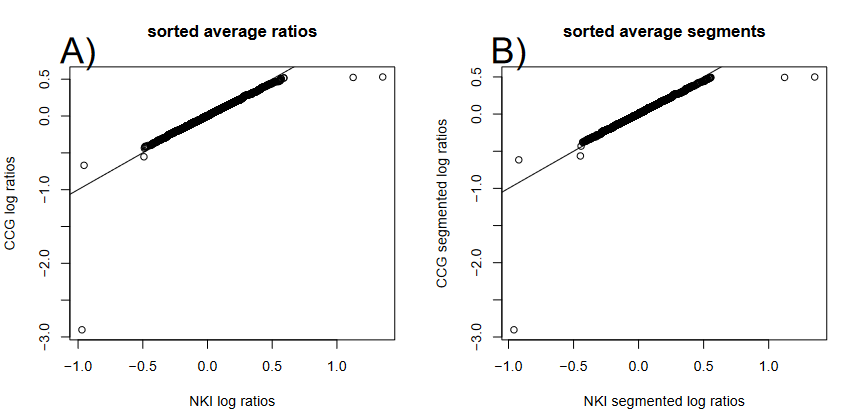

Supplementary Figure 2. Sorted average log ratios (A) and segmented log ratios (B) of both centers. The diagonal line represents x=y.

We plotted the distributions of the log ratios and segmented log ratios in Supplemental Figure 3. The distributions were very similar.

Supplementary Figure 3. Distribution of (segmented) log ratios of 47 samples sequenced at NKI and CCG.

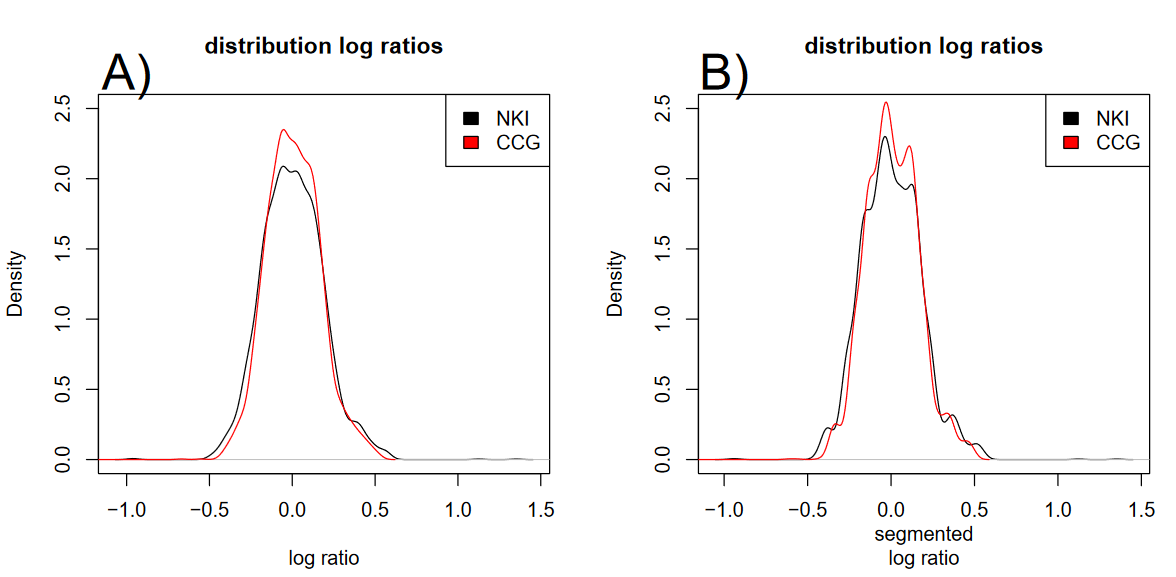

Supplementary Figure 3. Distribution of log ratios (A) and segmented log ratios (B) of 47 samples that were sequenced at NKI and CCG.

Given that the obtained copy number aberration profiles appeared very similar, specifically the segmented ratios which are used for classification, we classified the 47 samples and cross tabulated the results in Supplementary Table 1. 46/47 samples (98%) showed the same BRCA1-like classification, see Supplementary Table 1.

Supplemental Table 1.

|  |  | CCG |  |
| --- | --- | --- | --- |
|  |  | Not *BRCA1*-like | *BRCA1*-like |
| NKI | Not *BRCA1*-like | 23 | 1 |
|  | *BRCA1*-like | 0 | 23 |

Supplementary Table 1. *BRCA1*-like classification of 47 samples sequenced at NKI and CCG.

We plotted the copy number aberration profile (Supplementary Figure 4) of the discordant sample and observed both in the visual assessment, and confirmed by the density plot (Supplementary Figure 5), that the CCG-sequenced profile has reduced amplitude. Although the overall validation succeeded, there is residual experimental variation, despite the profile passing quality control.

Supplemental Figure 4. Copy number profile of discordant sample sequenced at NKI and at CCG.

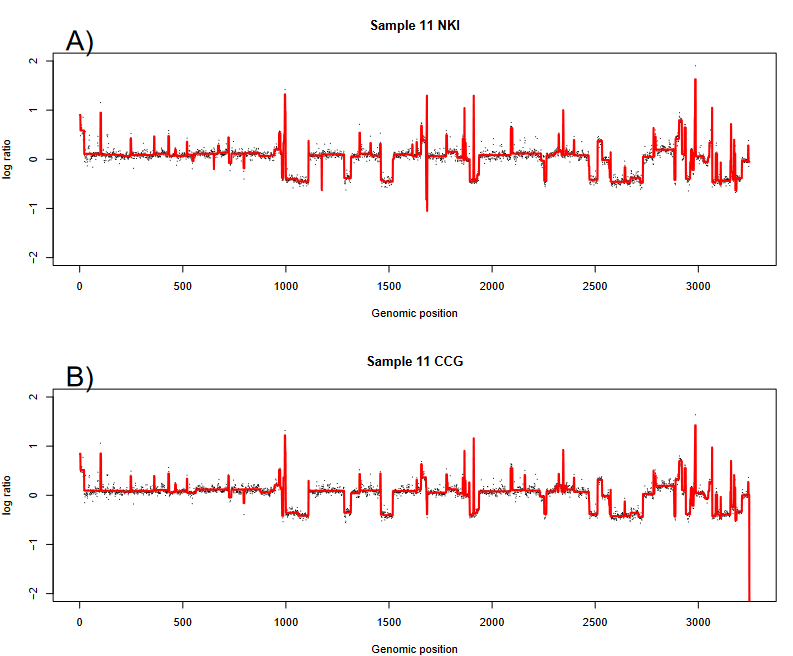

Supplemental Figure 5. Copy number aberration profile of the sample that is discordant between NKI (A) and CCG (B).

Supplementary Figure 5. Density plot of the sample that is discordant between NKI and CCG.

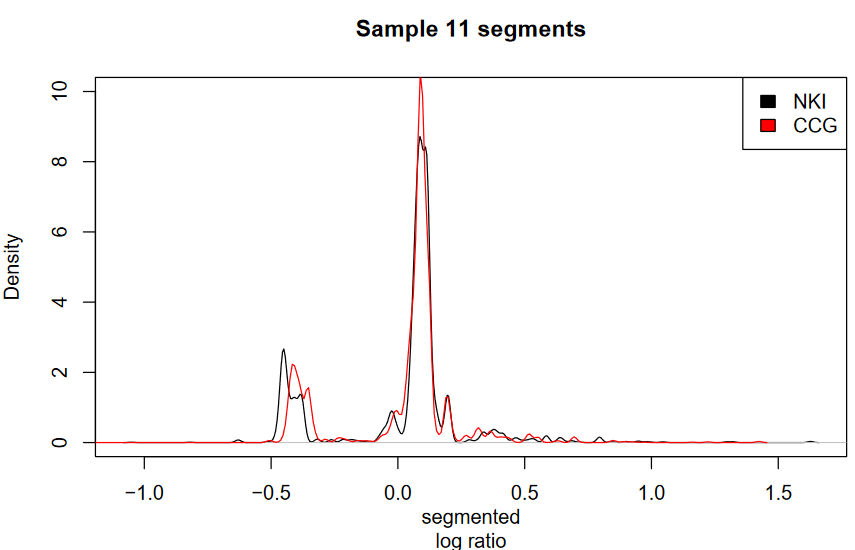

Supplementary Figure 5. Density plot of the segmented data of the discordant sample between NKI and CCG. Reduced amplitude for the CCG sample is observed, e.g. at the peak around -0.5 and in the values > 0.25.

*Additional Survival analysis*

Supplementary Figure 6. Kaplan-Meier curves of PFS and OS comparing olaparib+bevacizumab vs. placebo in all combination of mutation status and *BRCA*-like status.

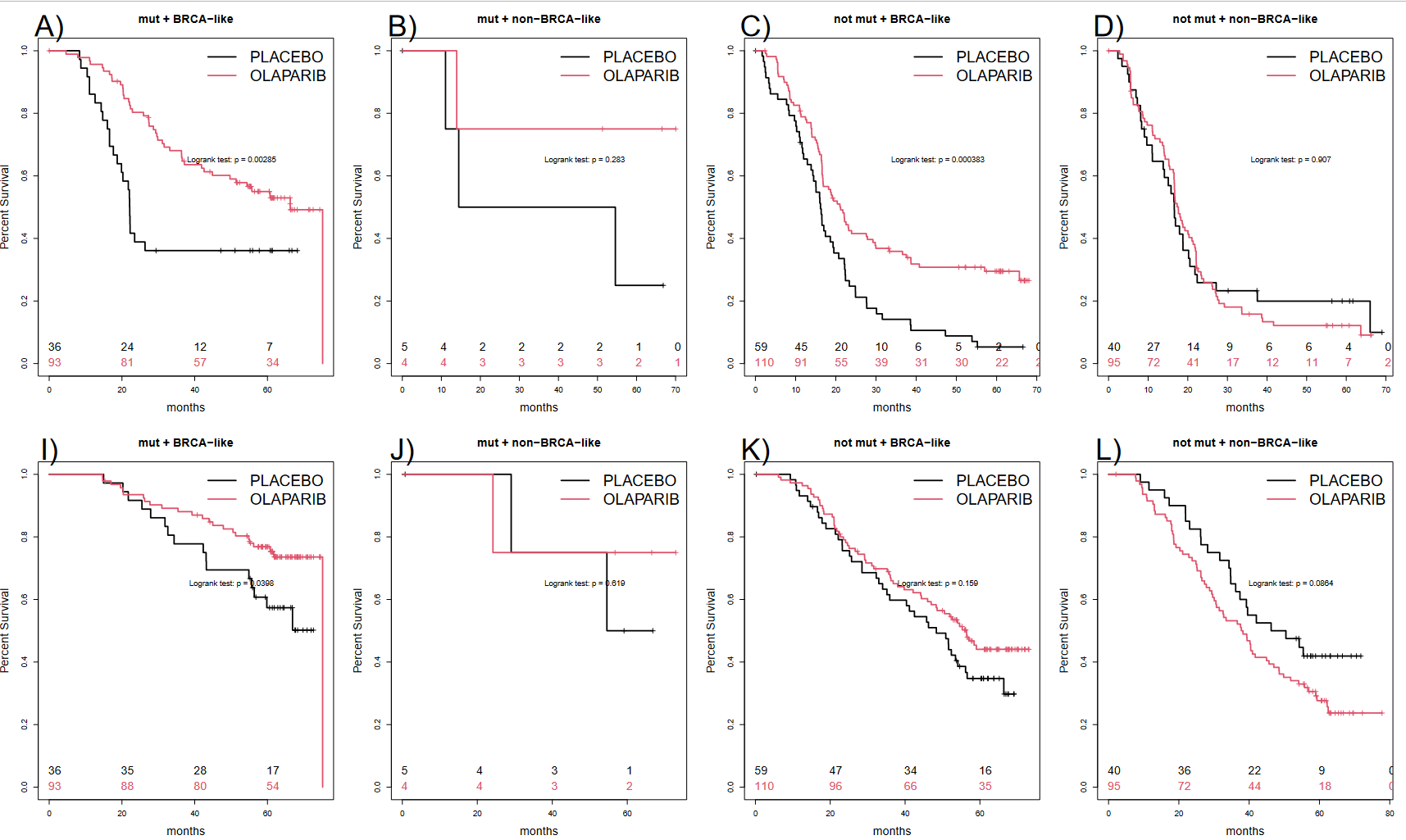

Supplementary Figure 6. Kaplan-Meier curves of PFS of olaparib + bevacizumab vs. placebo in patients with a mutation and *BRCA*-like tumor (A), with a mutation but non-*BRCA*-like tumor (B), without mutation and *BRCA*-like tumor (C), without mutation and non-*BRCA*-like tumor (D). Kaplan-Meier curves of OS of olaparib + bevacizumab vs. placebo in patients with a mutation and BRCA-like tumor (E), with a mutation but non-*BRCA*-like tumor (F), without mutation and *BRCA*-like tumor (G), without mutation and non-*BRCA*-like tumor (H). The hazard ratios are presented in Supplementary Table 2.

Supplementary Table 2. Hazard ratios and median survival of the Kaplan-Meier curves presented in Supplementary Figure 6.

| PFS |  |  |  |  |  |  |  |
| --- | --- | --- | --- | --- | --- | --- | --- |
|  |  |  | Median Survival | Treatment | HR | 95%CI | p |
|  | Mutation | BRCA-like |  |  |  |  |  |
|  | Yes | Yes | 22.1 | Placebo | 1 |  |  |
|  |  |  | 66.3 | Olaparib | 0.47 | 0.28-0.78 | < 0.01 |
|  | Yes | No | 34.5 | Placebo | 1 |  |  |
|  |  |  | not reached | Olaparib | 0.31 | 0.03-2.98 | 0.31 |
|  | No | Yes | 16.2 | Placebo | 1 |  |  |
|  |  |  | 21.2 | Olaparib | 0.53 | 0.38-0.76 | < 0.01 |
|  | No | No | 16.6 | Placebo | 1 |  |  |
|  |  |  | 17.6 | Olaparib | 1.03 | 0.68-1.55 | 0.90 |
| OS |  |  |  |  |  |  |  |
|  |  |  | Median Survival | Treatment | HR | 95%CI | p |
|  | Yes | Yes | not reached | Placebo | 1 |  |  |
|  |  |  | 75.2 | Olaparib | 0.52 | 0.27-0.98 | 0.04 |
|  | Yes | No | 54.6 | Placebo | 1 |  |  |
|  |  |  | not reached | Olaparib | 0.55 | 0.05-6.08 | 0.62 |
|  | No | Yes | 48.4 | Placebo | 1 |  |  |
|  |  |  | 56.4 | Olaparib | 0.75 | 0.5-1.12 | 0.16 |
|  | No | No | 48.4 | Placebo | 1 |  |  |
|  |  |  | 38.0 | Olaparib | 1.51 | 0.94-2.42 | 0.09 |

Supplementary Figure 7. Flow diagram of classification of samples by both tests.

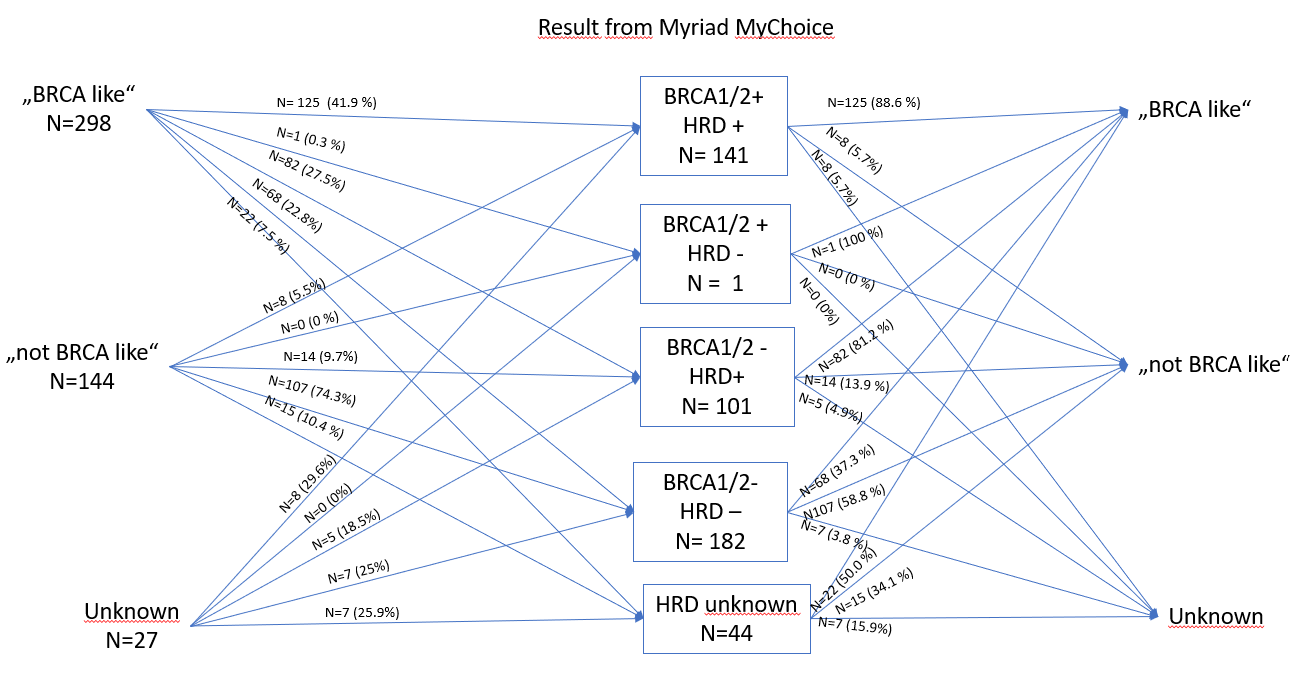

Supplementary Figure 7. Flow diagram of classification of samples by both tests. HRD = Homologous Recombination Deficient by Myriad MyChoice.
